# Continuous Estimation of Achilles Tendon Loading in Rupture Patients Using a Single Boot-Mounted Accelerometer

**DOI:** 10.64898/2026.03.10.26348070

**Authors:** Stanton Godshall, Lorraine AT Boakye, Eni Halilaj, Casey Jo Humbyrd, Josh R Baxter

## Abstract

**Objective:** Achilles tendon ruptures lead to long-term structural and functional deficits. Prior research that sought to identify optimal rehabilitation techniques was fundamentally limited by the inability to continuously monitor Achilles tendon loading during rehabilitation. Our objective was to develop a data-driven model that predicts per-step peak Achilles tendon loading from only a single, boot-mounted accelerometer.

**Methods:** Nineteen patients recovering from an acute Achilles tendon rupture completed in-lab walking trials while wearing an instrumented immobilizing boot. A boot-mounted inertial measurement unit provided acceleration signals used for prediction, while a force-sensing insole provided ground truth tendon-loading data through a validated ankle moment balance. We developed a stance-detection algorithm, as well as a personalized one-dimensional convolutional neural network (1D-CNN) to estimate per-step peak Achilles tendon load. Our training framework incorporated a small patient-specific personalization sample and was evaluated on held-out steps.

**Results:** The stance detection algorithm identified stance phases with 99.8% precision and mean timing errors of 27.3 ms for heel strike and 61.9 ms for toe-off. The CNN estimated per-step peak Achilles tendon load with a mean absolute error of 0.14 bodyweights (R^2^=0.68) across rupture patients.

**Conclusion:** Continuous, objective estimation of Achilles tendon loading during early rehabilitation is feasible using a single, boot-mounted accelerometer. Model errors were small (9%) relative to the wide range of tendon loading exhibited during immobilizing boot walking. Our proposed approach enables clinicians to continuously monitor mechanical loading during a previously unobservable rehabilitation period and provides a foundation for personalized rehabilitation guidance after Achilles rupture.

## Introduction

Achilles tendon ruptures are common injuries with an incidence of 7.8-41.7 occurrences per 100,000 person-years [1], [2], [3], [4] and have been increasing in frequency across North America and Europe [1], [5], [6], [7]. Studies show that patients suffer functional deficits that last at least 7 to 11 years after the injury [8], [9], and that nearly 20% of patients do not return to their previous level of physical activity [10]. Patients experience long-term structural alterations following rupture, including thicker, elongated, and less stiff tendons with compromised tissue composition [11], [12], [13] despite advances in clinical treatment [14], [15], [16]. These structural changes are closely linked to persistent functional deficits [17], [18], [19], [20], [21], suggesting that clinical strategies that improve tendon healing are likely to mitigate long-term functional deficits and ultimately improve clinical outcomes [22]. Rehabilitation protocols that protect the healing Achilles tendon while promoting regeneration are therefore a critical target for improving outcomes after Achilles rupture.

The Achilles tendon is mechanosensitive [23], and appropriate mechanical loading is thought to promote healing and regeneration [24]. However, following Achilles rupture, excessive loading may contribute to structural degradation like elongation or rerupture [22]. Randomized trials have directly tested how different rehabilitation protocols influence outcomes after Achilles rupture [9], [25], [26], [27], [28]. A guiding premise of these approaches is that rehabilitation protocols alter the mechanical environment of the healing tendon. Despite this, a critical limitation of prior rehabilitation research is that Achilles tendon loading has not been measured directly once patients leave the clinic. Instead, most studies infer loading from prescribed rehabilitation protocols [25], [26], [27], [29], implicitly assuming patient adherence, or rely on subjective self-report to capture mechanical exposure [30]. Therefore, true variations in tendon loading across patients and time remain unknown, which may explain why randomized trials have struggled to identify optimal rehabilitation strategies.

Wearable sensing offers a promising approach to address this measurement gap by continuously monitoring physiological signals outside of laboratory or clinical settings [31], [32]. Inertial measurement units (IMUs) are ubiquitous for collecting acceleration and angular velocity data that has been applied to the estimation of joint-level or tissue-level loading [33], [34], [35] but have not yet been applied to patients recovering from Achilles tendon ruptures. These patients present a distinct sensing challenge due to highly conservative gaits during early rehabilitation [36] and the complex constraints that immobilizing boots impose on Achilles tendon loading [37].

Our prior work demonstrated proof-of-concept in healthy adults by using IMU-derived features and a least absolute shrinkage and selection operator (LASSO) regression model to estimate peak Achilles tendon loading [38]. That study was limited because it excluded steps with peak loads below 0.5 bodyweights from analysis, a range that is common during rehabilitation after rupture [39]. Further, it relied on force-sensing insoles to segment IMU data into steps prior to the extraction of IMU-derived features, preventing deployment as a stand-alone, single-sensor solution.

Our engineering objective was to develop a load-estimation pipeline that is accurate and deployable in clinical practice. Commercially available wearable sensors can record for extended periods when configured to measure accelerations alone at modest sampling rates (e.g., 50 Hz), enabling monitoring between clinical visits with minimal burden. Accordingly, we aimed to estimate per-step peak Achilles tendon loading in Achilles rupture patients using only a single boot-mounted accelerometer sampled at 50 Hz. To achieve this, we combined a rule-based stance detection algorithm with a one-dimensional convolutional neural network (1D-CNN) to generate per-step peak Achilles tendon load estimates from accelerometer data alone in Achilles rupture patients. Peak Achilles tendon loads in our rupture cohort spanned 0.2-1.69 bodyweights during walking in an immobilizing boot (**Figure 7**). Since monitoring aims to resolve differences within this loading range, we defined our success criterion as a mean absolute error below 0.25 bodyweights (∼15% of the observed range) prior to model evaluation.

## Methods

### Participants

We recruited 19 participants (16 male, age: 35 ± 6 years, BMI: 26.7 ± 8.4 kg/m^2^) who received treatment for an acute Achilles tendon rupture. All participants were managed by one of four fellowship-trained orthopaedic surgeons. Most of the participants (17/19) opted for surgical repair. Participants visited the Human Motion Lab at the University of Pennsylvania 7.3 ± 2.3 weeks after treatment initiation, determined by the clinical visit where participants are weight-bearing in an immobilizing boot. Participants were eligible if they were aged 18-60 years, did not smoke, and did not use steroids in the last 6 months. All participants provided written informed consent prior to any study activities that were approved by the University of Pennsylvania Institutional Review Board (IRB: 850302).

### Healthy Controls

We leveraged a healthy control dataset that we previously collected [38] to develop, train, and conduct preliminary testing of our models. Briefly, 10 healthy controls (7 male, age: 25 ± 2.4 years, BMI: 23.9 ± 6.6 kg/m^2^) completed eight-meter walks in a standard immobilizing boot at various speeds, gait patterns, and wedge constraints. Data from healthy controls were collected with the same wearable sensing system as is deployed in the present study.

### Data Collection

During lab visits, participants completed approximately eight eight-meter walking trials through our laboratory space in their clinician-approved immobilizing boots. Crutches were not used to support weight-bearing on their injured leg. Participants walked at a self-selected walking speed. Brief pauses were allowed between each trial to prevent fatigue and recording only occurred during straight line walking. This procedure generated 1146 total steps across all rupture participants.

### Instrumentation

Immobilizing boots were instrumented with a commercially available IMU (Opal, APDM, Portland, OR) rigidly strapped to the proximal lateral portion of the immobilizing boot. The IMU was configured to measure accelerations with a measurement range of 16 g at 100 Hz. Additionally, we placed a three-part force-sensing insole (Loadsol, Novel, St. Paul, MN) inside the immobilizing boot according to our previously published protocol [37], [40] to estimate Achilles tendon loading during immobilizing boot ambulation (**Figure 1**).

**Figure 1.**
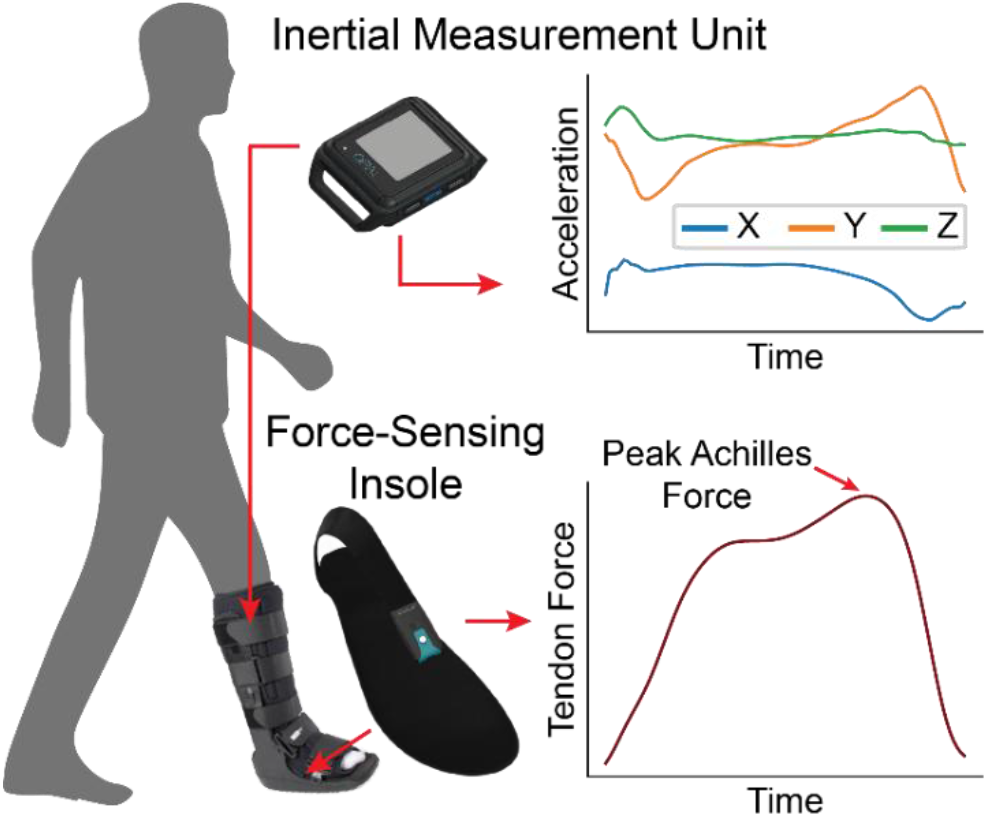
Sensor Placement and Output. An inertial measurement unit measures accelerations while a force-sensing insole measures tendon force.

### Stance Detection Algorithm

All analysis was performed in Python using custom-written scripts that leverage scientific computing libraries (SciPy signal processing module) [41]. We developed our stance detection algorithm on the data that were previously collected from the 10 healthy controls to reduce the risk of overfitting. Our algorithm identifies periods of flat accelerations that coincide with periods when the boot is on the ground. This contrasts with other techniques that rely on heel-strike or toe-off features or gyroscope data [42], [43]. These techniques are inapplicable because heel strike and toe off are not well defined in a walking boot. We defined these flat regions by calculating a sliding root-means-square (RMS) of 0.24-second-long windows of the acceleration magnitude (**Figure 2A and B**). We then identified regions where the sliding RMS is below the 40th percentile for at least 0.18 seconds (**Figure 2C**). To improve precision, we automatically fill gaps in flatness that are less than 0.1 seconds long. Additionally, if the signal climbs to 110% of its average value at either end of the flat region, we shrink the region until both ends are below 103% of the average value. Finally, we identify heel strike and toe off by identifying the peaks in acceleration magnitude before and after the flat region, respectively (**Figure 2D**). We require these peaks to have a magnitude at least 110% of the signal’s average value. We search for peaks in an adaptive window that has length between 35 and 50% of the duration of the flat region. To validate our stance detection algorithm, we compared it to our instrumented insole-derived heel strike and toe off. Precision is defined as the portion of stances that occur during true stances. Timing error is defined as the time between the algorithm-defined gait event and the moment where plantar force passes a predefined 50N threshold.

**Figure 2.**
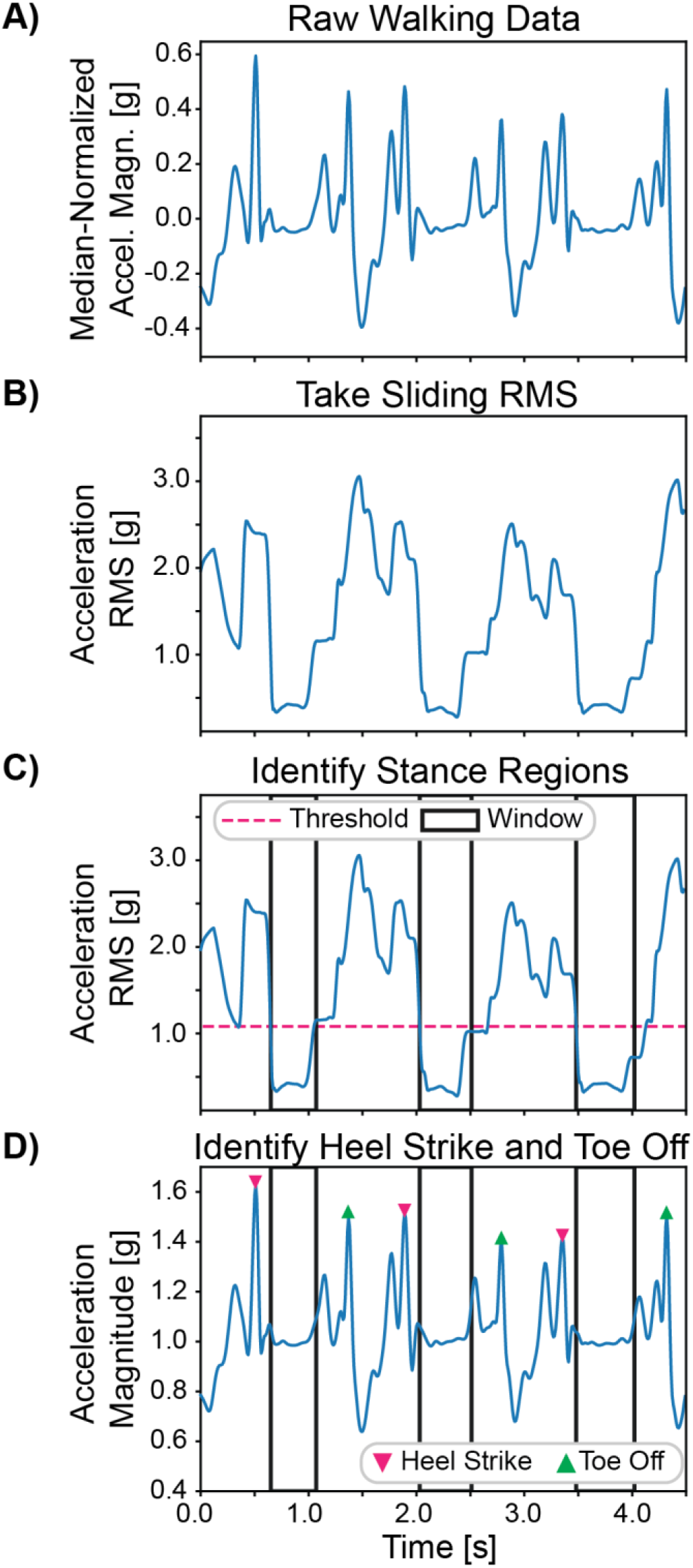
Stance Detection Algorithm. Stance is identified solely from acceleration data. A) Raw acceleration data from 3 consecutive steps. B) The sliding root-mean-square (RMS) of acceleration quantifies flatness. C) Flat regions are defined using the sliding RMS. D) Heel strike and toe off are identified as peaks that precede and succeed the flat region.

### 1D-CNN Training

We developed a one-dimensional convolutional neural network (1D-CNN, **Figure 3**) to estimate peak Achilles tendon loads on a per-step basis from triaxial accelerometer signals. Models were trained in batches of 64 steps. Each convolutional layer was followed by a rectified linear unit (ReLU) activation and a max-pooling operation with a size of 2. Inputs were normalized using statistics from the training data only. After flattening and concatenating the metadata, batch normalization and 50% dropout were applied prior to the fully connected layers and regression to our target scalar output. Models were trained with L1 Loss up to 20 epochs without early stopping and optimized with Adam [44]. Models were implemented and trained using PyTorch [45].

**Figure 3.**
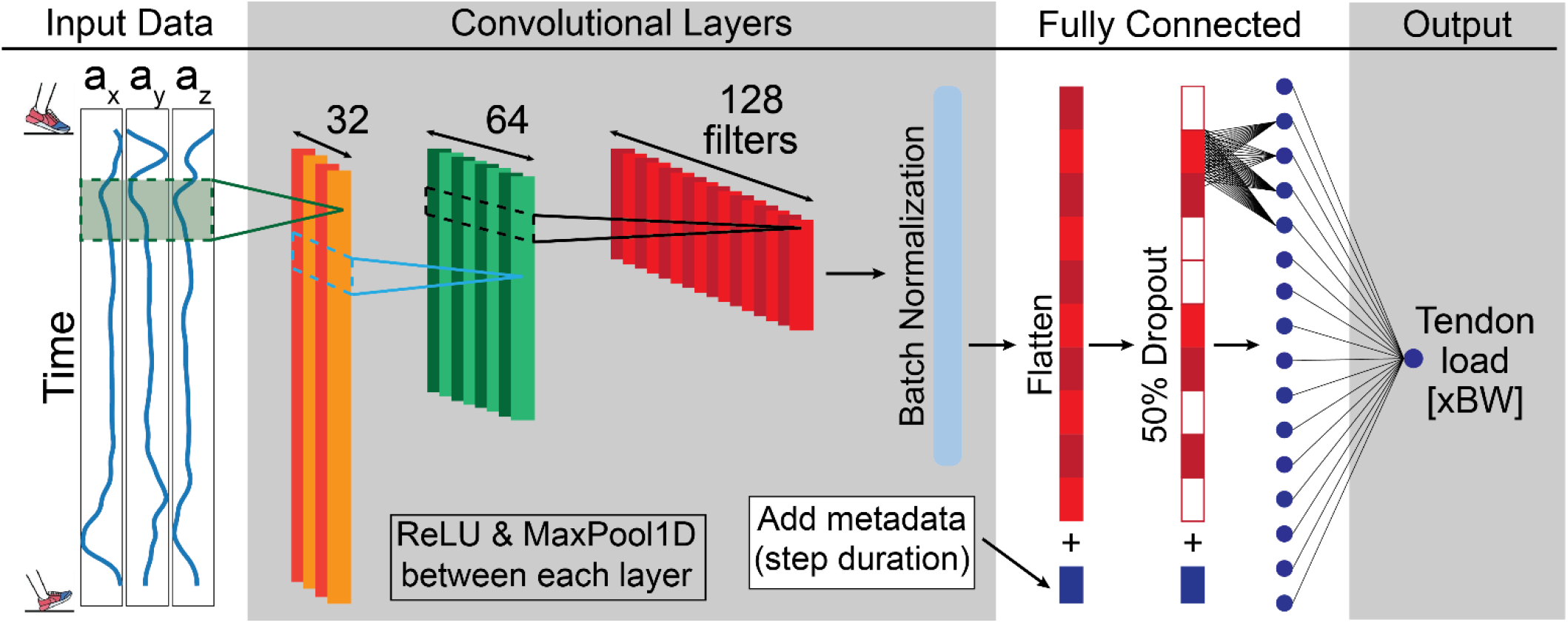
Sample 1D-CNN Architecture. Resampled triaxial acceleration time series are processed through a stack of 1D convolutional layers with ReLU activation and max pooling. Batch normalization is applied to the convolutional features. The resulting features are flattened and concatenated with step duration metadata. Dropout is applied at 50% before fully connected layers that regress to a single parameter, peak tendon load.

Triaxial acceleration time series data were downsampled to 50 Hz by decimation and subsequently resampled to 100 samples prior to CNN input. We used this sampling approach to simulate our deployment scenario where a commercially available accelerometer (AX6, Axivity Ltd, New Castle, UK) sampling at 50 Hz captures ∼35 days of continuous data – completely capturing patient activity between ∼4-week patient visit spacing. Following convolutional layers and flattening, the step duration in seconds is appended as scalar metadata. Thus, this algorithm operates solely with inputs derived from a single boot-mounted accelerometer sampling at 50 Hz. Labels were generated by identifying the peak Achilles tendon force during the identified step region. Insole data was filtered with a 4^th^-order Butterworth filter with a cutoff frequency of 10 Hz and used to calculate ground truth Achilles tendon loads using our validated algorithm [40].

We followed a personalized, per-participant model training framework (**Figure 4**). For each rupture participant, 50% of that participant’s steps were intentionally included in training and the other 50% was held out for model evaluation. In addition to the participant’s 50% of the data, the training dataset also included all the data from the 18 remaining patients and 10 healthy controls. We divided the remaining rupture participants using five-fold cross-validation to select hyperparameters. We did not perform hyperparameter selection on healthy control data to bias the model towards better performance on rupture participants. Within each validation fold, 50% of the steps from the participants in the validation set were added to training to mimic the personalization approach of the outer loop. This procedure yielded a personalized model for each participant, with performance evaluated exclusively on each participant’s held-out steps. This approach improved accuracy in our prior work [38] and is feasible given the small model size and our ability to capture patient loading data at each clinical visit. We included all data from the healthy controls (7952 steps) to expose the model to a greater diversity of gait strategies [38]. Healthy controls were similarly evaluated with the personalized framework; however, their training did not include rupture patient data and did not use 5-fold cross-validation.

**Figure 4.**
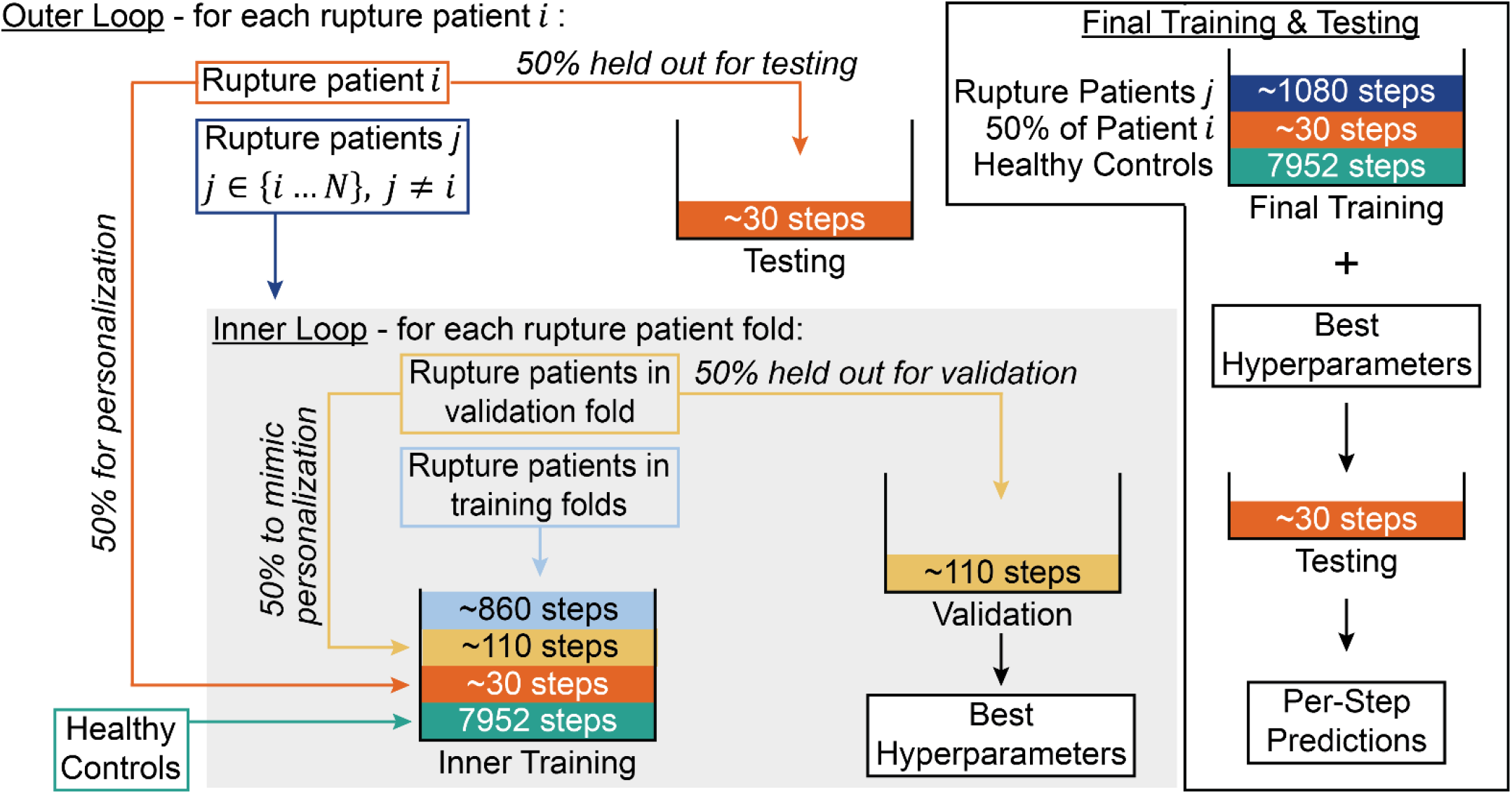
Personalized Training Procedure. To expose the 1D-CNN to patient-specific gait strategies, our model incorporates a sample of each testing patients data into training. This procedure is repeated during hyperparameter selection, where validation participants contribute a sample of their data into training. Healthy control data is included in training to diversify the model’s exposure.

Hyperparameters were selected using grid search within the inner cross-validation loop by identifying the minimum mean validation loss across validation folds. The hyperparameter space included varying convolutional stacks with options [32,64], [32,64,64], and [32,64,128], kernel sizes 3 and 5, fully connected layers with 16 and 32 units, and learning rates of 10^-3^ and 10^-4^. Upon determination of the best hyperparameters, final training commenced with all healthy controls, all rupture patients that are not held out, and 50% of the steps from the evaluation subject.

### Model Evaluation

Our target for clinical impact was a model that estimates peak Achilles tendon load with a mean absolute error below 0.25 bodyweights. We further evaluated model performance without using the personalized framework to assess performance when personalization is not viable. We systematically shrunk the personalization sample size by reducing its size by 25% increments, down to 0 total personalization steps. Training the model with a personalization sample size of 0 is equivalent to leave-one-subject-out cross-validation. In this analysis, we utilized the hyperparameters that were selected in our original test.

## Results

The stance detection algorithm identified stance phases from accelerometer data during immobilizing boot walking in Achilles rupture patients with 99.8% precision (**Figure 5A and B**). Mean absolute timing error was 27.3 ms at heel strike and 61.9 ms at toe off relative to force-sensing insole-derived timing (**Figure 5C**).

**Figure 5.**
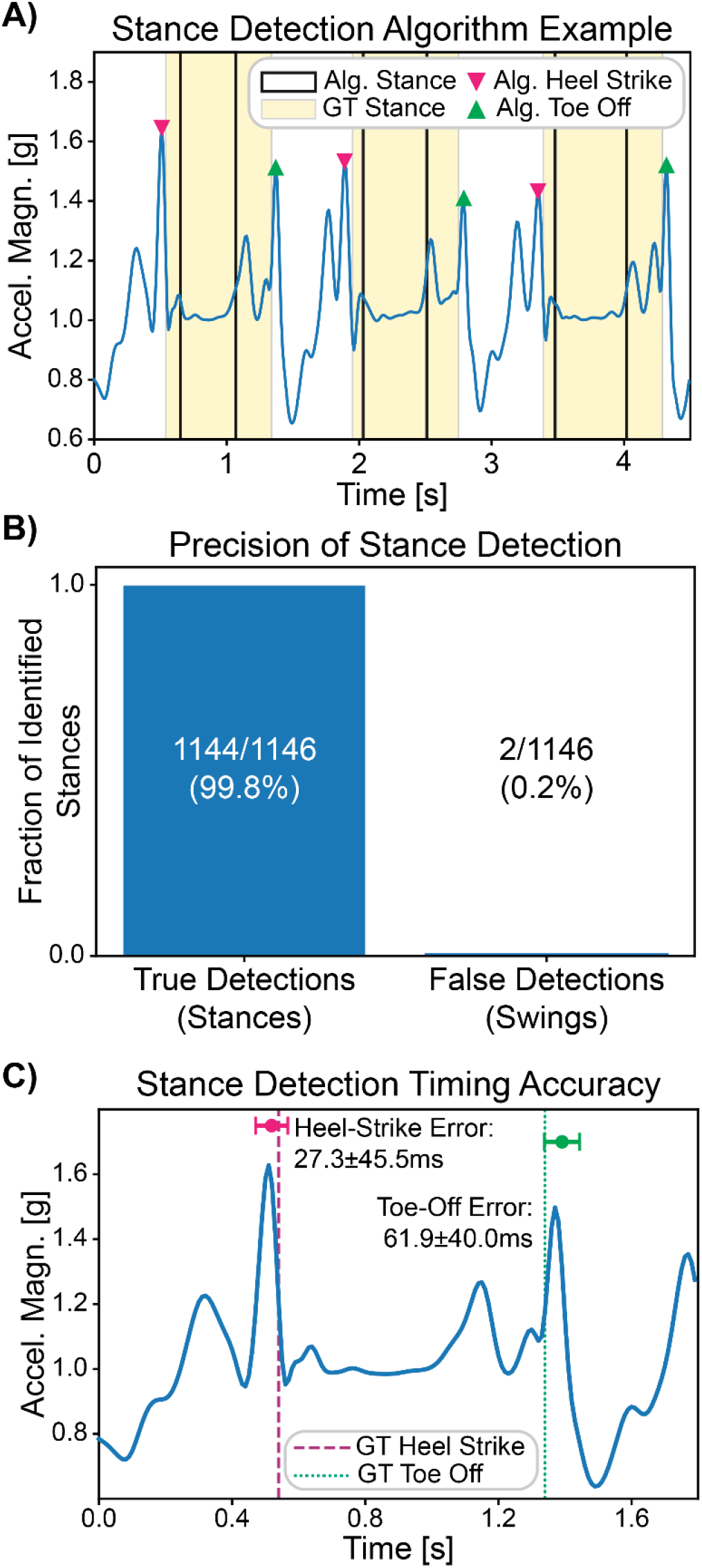
Stance Detection Algorithm Results. In Achilles rupture patients, the stance detection algorithm accurately parses stance phases used as input to the 1D-CNN. A) an example of three consecutive steps shows algorithm performance. B) Of identified stance phases, 99.8% are true stance phases, while 0.2% are incorrectly identified swings. C) Relative to force-sensing insole-derived timing, heel-strike and toe-off are identified with 27.3 and 61.9 ms of error, respectively.

The personalized 1D-CNN estimated per-step peak Achilles tendon load with a mean absolute error (MAE) of 0.14 bodyweights across patients and an R^2^ of 0.68. Similarly, per-step peak Achilles tendon load was estimated with a MAE of 0.24 bodyweights and an R^2^ of 0.87 across healthy controls (**Figure 6**).

**Figure 6.**
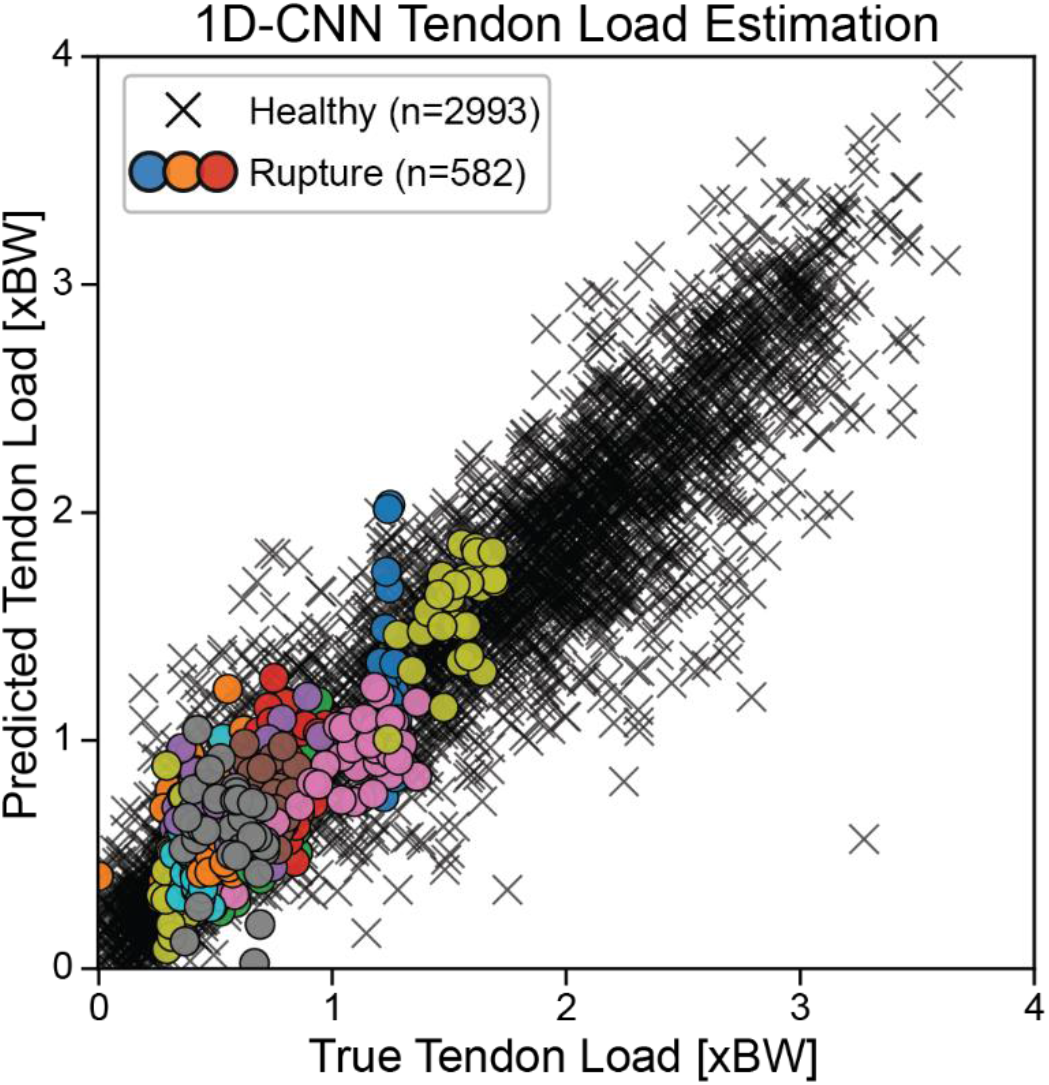
1D-CNN Load Estimation Results. Each step is plotted as predicted peak Achilles tendon load versus true peak Achilles tendon load. Each step for Achilles rupture patients is plotted with a patient-color-coded circle, while healthy control steps are plotted with a black X.

Model errors varied across patients, with a per-patient minimum absolute error of 0.01 bodyweights and a maximum absolute error of 0.19 bodyweights (**Figure 7A**). Signed errors demonstrated a moderate correlation with the true peak Achilles tendon load (r = -0.47, p=0.04), where patients with lower loads tended to be overestimated and patients with higher loads tended to be underestimated (**Figure 7B**).

**Figure 7.**
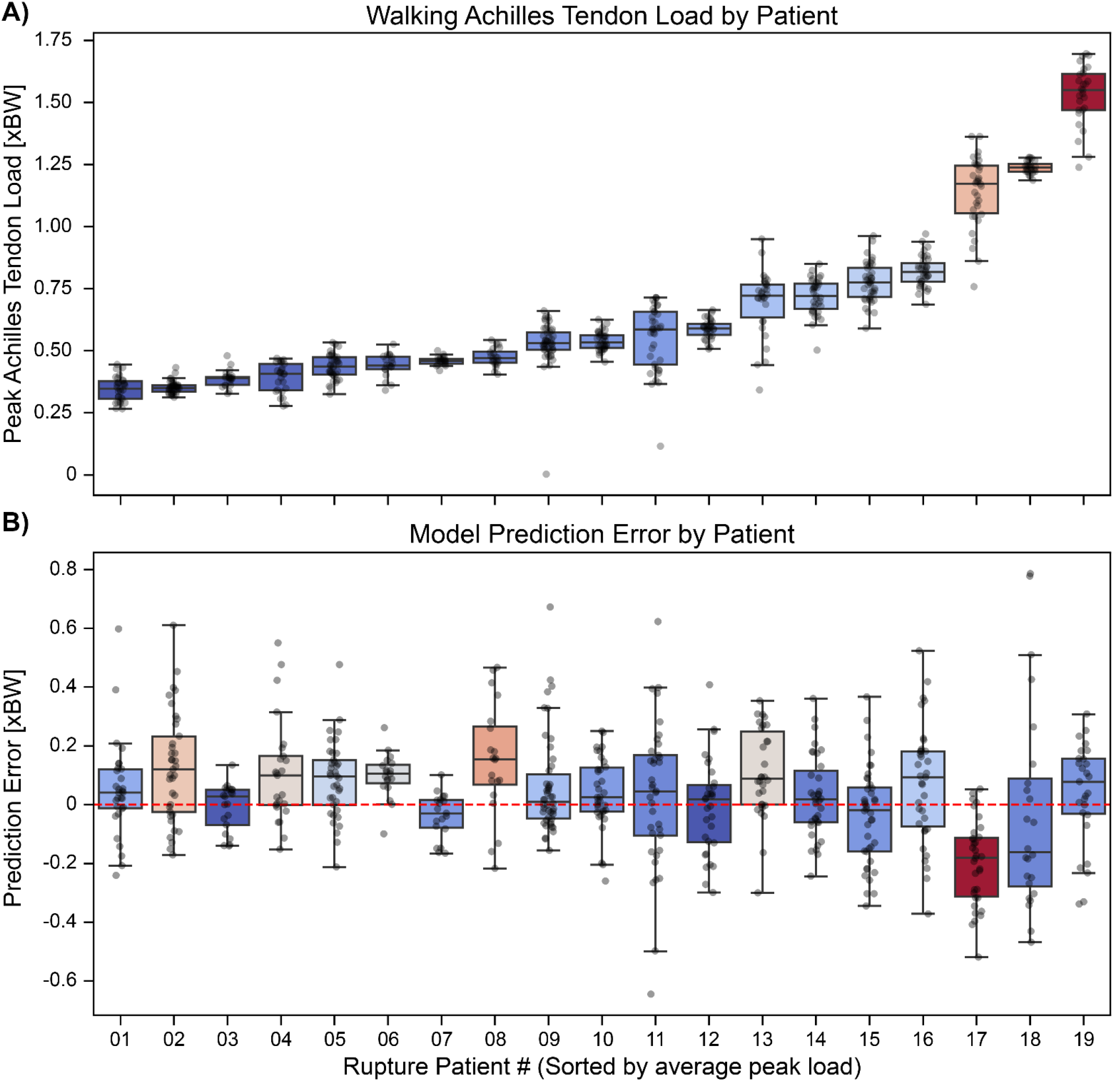
Distribution of Errors by Peak Load and Patient. Patients are ordered on the x-axis according to their peak Achilles tendon load. A) The true peak tendon load is plotted per step per patient. B) The prediction error for each step is plotted per patient. Boxes show the median and interquartile range. The color encodes the absolute value of the y-axis.

A sensitivity analysis that varied the size of the personalization sample revealed that model performance declined as fewer participant-specific steps were included. Under full leave-one-subject-out cross-validation, model performance decreased to an average MAE of 0.29 bodyweights. The range of absolute errors increased to a minimum of 0.09 bodyweights and a maximum of 0.71 bodyweights (**Figure 8**).

**Figure 8.**
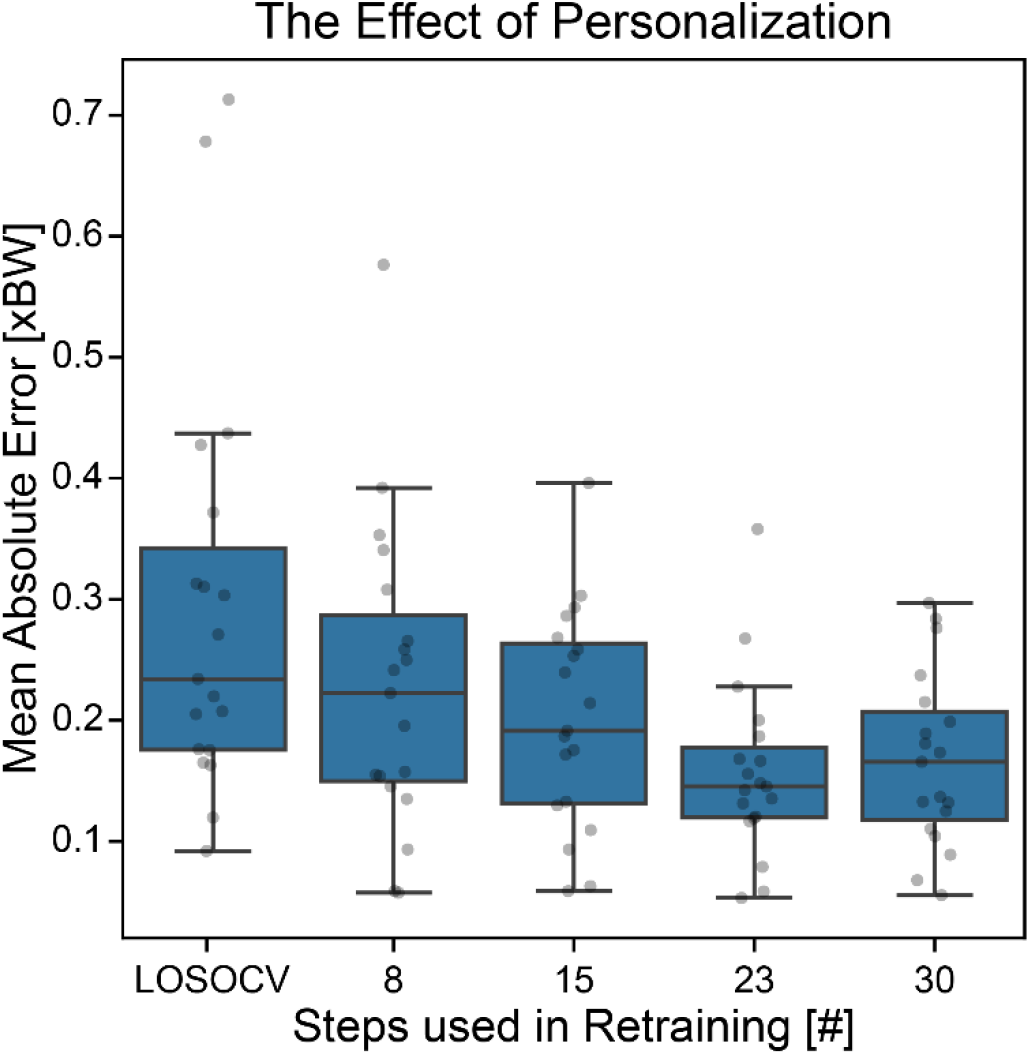
Sensitivity Analysis of Personalization Sample. Per-patient mean absolute error is plotted against the number of test-patient steps used in personalization. The zero personalization steps condition corresponds with leave-one-subject-out cross-validation (LOSOCV).

## Discussion

The goal of this study was to develop an algorithmic pipeline to convert periods of immobilizing boot walking into per-step peak Achilles tendon load estimates using only a single, boot-mounted accelerometer. We found that stance phases can be identified with 99.8% precision by targeting the acceleration flatness during stance. This step is critical because successful load estimation requires models that are trained and deployed on the correct distribution of data. Furthermore, we found that these stance phases can be reliably transformed into per-step peak Achilles tendon load estimates with a MAE of only 0.14 bodyweights across rupture patients. This shows that the model can identify trends in the accelerometer data that correspond to the loading that was required to produce the step. Taken together, these results support the notion that continuous load estimation is achievable with a single boot-mounted accelerometer in Achilles rupture patients.

This study addresses a fundamental limitation in prior Achilles rupture rehabilitation research by enabling continuous, objective measurement of Achilles tendon loading rather than relying on prescribed protocols or subjective self-report to capture true mechanical exposure. By estimating per-step tendon loading continuously, this approach provides a way to quantify the mechanical stimulus experienced by the healing tendon, enabling more precise associations between loading patterns and biological or clinical outcomes. Importantly, this capability is achieved using a low-burden, deployment-oriented sensing strategy. The algorithm was designed under the engineering constraint that only a single boot-mounted accelerometer sampled at 50 Hz would be available at deployment. This constraint is aligned with current wearable devices, which can record for over a month on a single charge with these settings. This allows for convenient sensor swapping at monthly clinical visits. Such a configuration supports extended monitoring between clinical visits with minimal patient and clinician burden, creating a practical opportunity for integrating objective tendon loading estimates into rehabilitation research, and possibly future rehabilitation protocols.

These results may give the impression that the load-estimation 1D-CNN is simultaneously inferior and superior at predicting peak Achilles tendon loads in healthy controls, as the average MAE was worse by 0.1 bodyweights, but the R^2^ was greater by 0.19. A visual inspection of the 1D-CNN performance clarifies this discrepancy (**Figure 6**). It appears that the difference in estimation quality reflects differences in our evaluation criteria rather than true model estimation quality. In rupture patients, the range of peak loads is greatly reduced, which tends to decrease MAE while lowering R^2^.

Importantly, model performance was consistent across patients despite substantial heterogeneity in self-selected loading patterns. Our results indicate that participants self-select different gait patterns that produce markedly different peak Achilles tendon loads. These Achilles loads range from 0.1 to 1.69 bodyweights when measured using our instrumented insole in this cohort (**Figure 7**). We expect that resolving Achilles tendon loading to ∼10% of this loading range is sufficient to accurately quantify loading progression throughout healing to potentially support decision making for accelerated rehabilitation. The model achieved a mean absolute error of 0.14 bodyweights, corresponding to ∼9% of the observed loading range. Our mean absolute error of 0.14 bodyweights is well below the differences in tendon loading during the clinical progression of Achilles tendon exercises like seated heel raise, squatting, standing 2-leg heel raise, and standing 1-leg heel raise [39]. This supports our paradigm of monitoring tendon loading during protected weight-bearing by framing model accuracy in the context of clinical loading progression. Overall, our model presents a modest range of errors, with the worst patient only having 0.18 bodyweights more error on average than the model’s best predicted patient. Signed errors were moderately correlated with true peak load magnitude, indicating a tendency towards overestimation at low loads and underestimation at high loads. This pattern suggests that there may be signals not captured by the accelerometer that contribute to residual variance in peak tendon loading. This may cause our model to depress the true differences in tendon loading at a larger scale. However, the magnitude of this compression effect was small relative to the between-patient differences in tendon loading observed in this cohort, indicating that the model preserves clinically meaningful variability across patients.

Incorporating a small, subject-specific personalization sample improved accuracy relative to leave-one-subject-out cross-validation, consistent with our prior findings [38]. We interpret this gain as arising from increased exposure to patient-specific gait strategies rather than overfitting, as the personalization sample constitutes less than 1% of the total training data. When the personalization sample was removed, overall model accuracy decreased modestly, with an increase in average MAE of about 0.15 bodyweights. However, in a subset of participants, errors increased dramatically, with per-participant mean absolute errors reaching 0.71 bodyweights. This finding potentially supports the conclusion that personalization disproportionately benefits individuals with more atypical gait strategies, rather than uniformly improving performance across participants. We are thus optimistic that with a larger cohort, our model may perform better in leave-one-subject-out evaluations. From a deployment perspective, this approach remains feasible, as collecting a small personalization sample (<30 steps) during a clinical visit is practical and the burden of personalized models is minimal. Even without personalization, model performance remained sufficient to detect large differences in tendon loading between patients. This indicates that our personalized training framework improves accuracy but is not strictly required to capture clinically meaningful variability.

This study has several key limitations. First, the proposed load estimation pipeline was evaluated exclusively in a controlled, in-lab environment during early rehabilitation, and it has not yet been tested in more diverse walking contexts or as gait patterns continue to change throughout recovery. Model performance may therefore differ in earlier or later rehabilitation periods or in real-world environments. Second, Achilles tendon loading was estimated with a force-sensing insole and a validated moment balance about the ankle, which represents the best current practice in clinical biomechanics but is not a direct measurement of tendon loading. Third, the present model estimates only peak tendon load per step and does not capture other potentially relevant loading behaviors such as impulse or rate of force development, which may also influence tendon healing or damage. Finally, this framework was evaluated only during walking, which is appropriate during immobilization but may not generalize to other activities performed during rehabilitation. Despite these limitations, the proposed approach provides a scalable method for continuously and objectively monitoring Achilles tendon loading during a clinically important and previously unobservable phase of recovery.

## Conclusion

We developed and validated a deployable wearable-sensor pipeline that estimates per-step peak Achilles tendon loading during immobilizing boot walking using a single, boot-mounted accelerometer in Achilles rupture patients. By combining stance detection with a personalized 1D-CNN, this framework enables continuous, objective monitoring of tendon loading during rehabilitation. Prior clinical studies that investigated the effects of rehabilitation strategies were limited to indirect or subjective measures of this important exposure. Our 1D-CNN model demonstrated clinically meaningful accuracy across participants and remained informative even without personalization. While future work is needed to extend this approach to real-world settings, this study establishes a scalable method to potentially support the identification of optimized rehabilitation protocols and ultimately data-driven, patient-specific guidance during Achilles tendon rehabilitation.

## Data Availability

All data produced in the present study are available upon reasonable request to the authors

## Acknowledgements

We thank Devyn Russo for assistance in patient recruitment and data collection and Michelle Kwon for assistance in healthy control recruitment and data collection.

## Data Availability

Data will be made available upon reasonable request.

